# Automated Digital Outreach with Human in the Loop is an Effective strategy for Recruitment and Electronic Patient Reported Outcomes: Results from All4IBD Multi-Centric Pragmatic Trial

**DOI:** 10.1101/2025.07.25.25331443

**Authors:** Stavros Stefanopoulos, Connie Chan, Morish Shah, Mounir Ibrahim, Jacqueline Bird, Drew Helmus, Catherine McLeod, Pamela Reyes Mercedes, Brian Baker, Lindsey Kaydo, Priya Patel, Sravya Kurra, Gaurav Narang, Shashank Garg, Sarthak Kakkar, Benjamin L. Cohen, Bruce E Sands, Maneesh Dave, Ashish Atreja, Maged Rizk

**Author notes:** Corresponding Author: Stavros Stefanopoulos, MD. Co-Senior Authors (Ashish Atreja, Maged Rizk). This is a clinical trial registered at ClinicalTrials.gov under ID: NCT04345393 titled Translating Scientific Evidence into Practice using Digital Medicine and Electronic Patient Reported Outcomes URL: https://clinicaltrials.gov/study/NCT04345393?id=NCT04345393&rank=1. Guarantor of the article: Maged Rizk, MD and Ashish Atreja, MD, MPH. Potential Competing Interest: -Dr. Atreja holds a patent for the RxUniverse (Prescription Universe) software platform that is licensed from Icahn School of Medicine at Mount Sinai to <u>Rx.Health</u>, Inc. (New York, NY) and acquired by Commure Inc. (MountainView, CA). Ashish Atreja owns stock in Commure, Inc and has recused himself from data analysis. - Sarthak Kakkar, Sravya Kurra, Garauv Narang, and Priya Patel are employees of Commure Inc. (Mountain View, CA) and own stock. They have recused themselves from data analysis.

## Abstract

**Objectives:** Patient recruitment is a critical factor for successful completion of a clinical trial. Decentralized trial recruitment allows potential subjects to be identified via the electronic health record (EHR) and approached through digital channels. We sought to understand the real-world effectiveness of decentralized trial recruitment leveraging electronic patient reported outcomes (ePROs) in patients with inflammatory bowel disease (IBD).

**Methods:** As part of a National Institutes of Health (NIH) funded multi-site clinical trial, we designed an EHR-integrated decentralized trial recruitment process to contact eligible patients at three tertiary IBD centers. We leveraged the Commure Engage/Rx.health (Mountain View, California) digital health formulary and automation engine integrated with Epic Systems (Epic Systems, Verona, WI) to digitally approach patients in an automated manner (with human-in-the loop) to complete the online enrollment process and monitor adherence with ePROs.

**Results:** Using digital outreach complemented by in-person engagement, we approached 6,687 eligible patients over 12 months and successfully enrolled 543 patients (mean age 40.7 ± 15.7 years, 63.3% females, 39.4% ulcerative colitis, 60.6% Crohn’s disease). 81.2% were recruited from bulk outreach methods. Gender (P<0.01), and race (P<0.01) were significantly associated with digital clinical trial enrollment. Patients were continually monitored with ePROs throughout the duration of the study.

**Conclusions:** This is one of the first studies to show the feasibility and successful recruitment of patients with IBD using automated digital outreach. We found a combination of outreach methods with human in the loop an effective strategy for clinical trial accrual. These findings suggest that ePROs can be successfully used in IBD centers to support treat-to-target strategy.

**Study Highlights:**

- What is Known:

- The most common reason for clinical trial failure is lack of accrual.
- Clinical trial recruitment strategies have remained largely unchanged despite technological advances.
- There is limited real world data around the use of electronic patient reported outcomes on a digital platform for patients with inflammatory bowel disease
- What is New Here

- ne of the first studies to employ a prospective decentralized recruitment approach for a multicenter clinical trial in inflammatory bowel disease.
- feasibility of an automated digital outreach for enrollment and continued monitoring using electronic patient reported outcomes

## INTRODUCTION

Participant recruitment is often the largest hurdle to overcome in the completion of a clinical trial. The National Institutes of Health (NIH) examined reasons for termination across all clinical trials occurring between 2007 - 2013 and found that insufficient accrual accounted for 57% of trials that ended prematurely.^1^ In a review of all Phase 3 interventional, two-arm, randomized control trials taking place in the United States between 2013-2014, 24% of the studies were discontinued citing low rates of enrollment as a significant impediment when compared to completed trials.^2^ In the current climate of scarce resources for medical research funding, the opportunity cost of each clinical trial failure and inefficiency represents a tremendous waste of potential to help the public. We have seen great advancements regarding the integration of technology into many aspects of medicine, however, these developments have yet to be fully realized in the field of clinical trial recruitment and patient assessment. National groups, such as the NIH and Trial Innovation Network, have begun to focus on improving the implementation of clinical trials.^3^ Some of these efforts have led to the idea of the decentralized clinical trial which the Decentralized Trials & Research Alliance (DTRA) defines as “A clinical trial utilizing technology, processes, and/or services that create the opportunity to reduce or eliminate the need for participants to physically visit a traditional research site.”^4^ This concept of reducing the physical presence of participants was further accelerated during the pandemic as studies began implementing decentralized processes. Decentralized clinical trials and electronic patient reported outcomes (ePROs) thus offer an exciting opportunity to bridge these gaps in clinical medicine and improve treat-to-target outcomes in patients with inflammatory bowel disease (IBD).

The concept of treat-to-target was first formalized in the Selecting Therapeutic Targets in Inflammatory Bowel Disease (STRIDE) guidelines and has since become the standard strategy in patients with IBD.^5^ We sought to understand the real-world effectiveness of decentralized trial recruitment leveraging ePROs in patients with IBD to augment the treat-to-target strategy. Our study examined how a decentralized recruitment and digital patient monitoring strategy through ePROs could be applied to a multi-institutional clinical trial at three academic medical centers (Mount Sinai, Cleveland Clinic, and University of California Davis) for patients with IBD. The eventual goal of the All4IBD pragmatic trial is to see if ePROs can be successfully used across IBD centers to proactively monitor patients’ symptoms and quality of life in line with the treat-to-target philosophy. This study is taking place over multiple years and in this manuscript, we present the successful pilot data of the decentralized recruitment process over a two-year process, while the ePRO monitoring is still ongoing at the time of publication.

## MATERIAL AND METHODS

We performed a NIH-funded multi-site trial focused on patients with IBD to first enroll patients via a decentralized recruitment process and then to establish baseline levels of disease control while implementing precision-matched interventions to improve outcomes using ePRO assessments at three tertiary IBD referral centers across the United States: Mount Sinai, University of California, Davis (UC Davis Health), and Cleveland Clinic. The trial was registered at clinicaltrials.gov with registration number NCT04345393. We utilized a centralized institutional review board (IRB) at all sites via a SMART IRB framework. A single centralized IRB protocol was created using Advarra services. Each site then reviewed the centralized IRB for approval at their institution which included ethical aspects of implementing and studying the intervention. A common protocol was submitted to Advarra Central IRB which addressed any potential conflicts of interest. The majority of communication throughout the study was performed utilizing a software platform developed by Commure Engage/Rx.health (Commure Inc, Mountainview, CA). We utilized this to facilitate the decentralized recruitment as well as ePRO monitoring. Subjects would enroll in communications options through the Commure Engage App Prescription Dashboard (Prescription Universe). Subjects would subsequently receive surveys via phone or email from the Prescription Universe.

Consistent with the pragmatic nature of trial, each site was able to customize its digital toolkit and leverage site-specific outreach, implementation and engagement processes. For example, language in the study protocol provided to patients was adjusted to accurately identify the site the patient would participate in however the content of the messages was the same across all sites. Each site also tailored the use of the Prescription Universe to better fit their needs. Specifically, UC Davis Health and Mount Sinai utilized Bulk Prescription for the initial outreach where Cleveland Clinic utilized the initial outreach via MyChart (Epic Systems) and then all subsequent patient interaction including enrollment via the Prescription Universe. At UC Davis Health, patients were only engaged who had documented in their electronic health record (EHR) that they were agreeable to being contacted for research. At Mount Sinai and Cleveland Clinic, interest in clinical research was not a data point that was captured prior to outreach for this project. The entirety of the study was conducted in English language only to patients at all sites.

Inclusion criteria was the same at each site and included: patients over the age of 18 with a documented IBD diagnosis managed by an IBD specialist at one of the three participating institutions, self-reported gaps in disease control identified through a screening questionnaire, willingness to comply with all study procedure as identified through a screening questionnaire, and access to necessary resources for participating in a technology-based intervention or willingness to get trained on digital resource stills. Exclusion criteria included patients with short bowel syndrome, multiple fistulae, history of bowel resection, stoma, or other GI conditions that make measurement of disease control not feasible via ePROs. Additionally, we excluded patients that had the presence of a condition or disease that in prohibited enrollment in digital monitoring including advanced dementia. Once patients were identified through digital query of existing registries or the EHR using Epic based search query language (SQL), we proceeded with a series of EHR-integrated decentralized recruitment processes as described below. Epic was used at all three sites regarding the prescreening process of patient charts. All other aspects of patient communication took place over the Prescription Universe dashboard or via telephone with coordinators at each site. Patient exclusion criteria were determined by first by a manual prescreening by research coordinators and then subsequently eligibility was confirmed by the study’s eligibility survey that the patients self-reported. Patients could consent then after these two rounds of eligibility screening. Those who became ineligible later during the study were opted out of the study if at any point they became ineligible; for example, if they had surgery during the study. Methods to identify and recruit research participants were split into three categories: Bulk Prescription, One-to-One prescription, and Text to Enroll. Regardless of outreach method though, patients who were ultimately enrolled in the trial experienced the same control and interventional period outlined in Figure 1. Human in the loop was also utilized at various stages and refers to any time the coordinator interacts with the subject such as via phone calls or face-to-face. This contrasts with automated communication through the Prescription Universe which would not be human in the loop.

**Figure 1.**
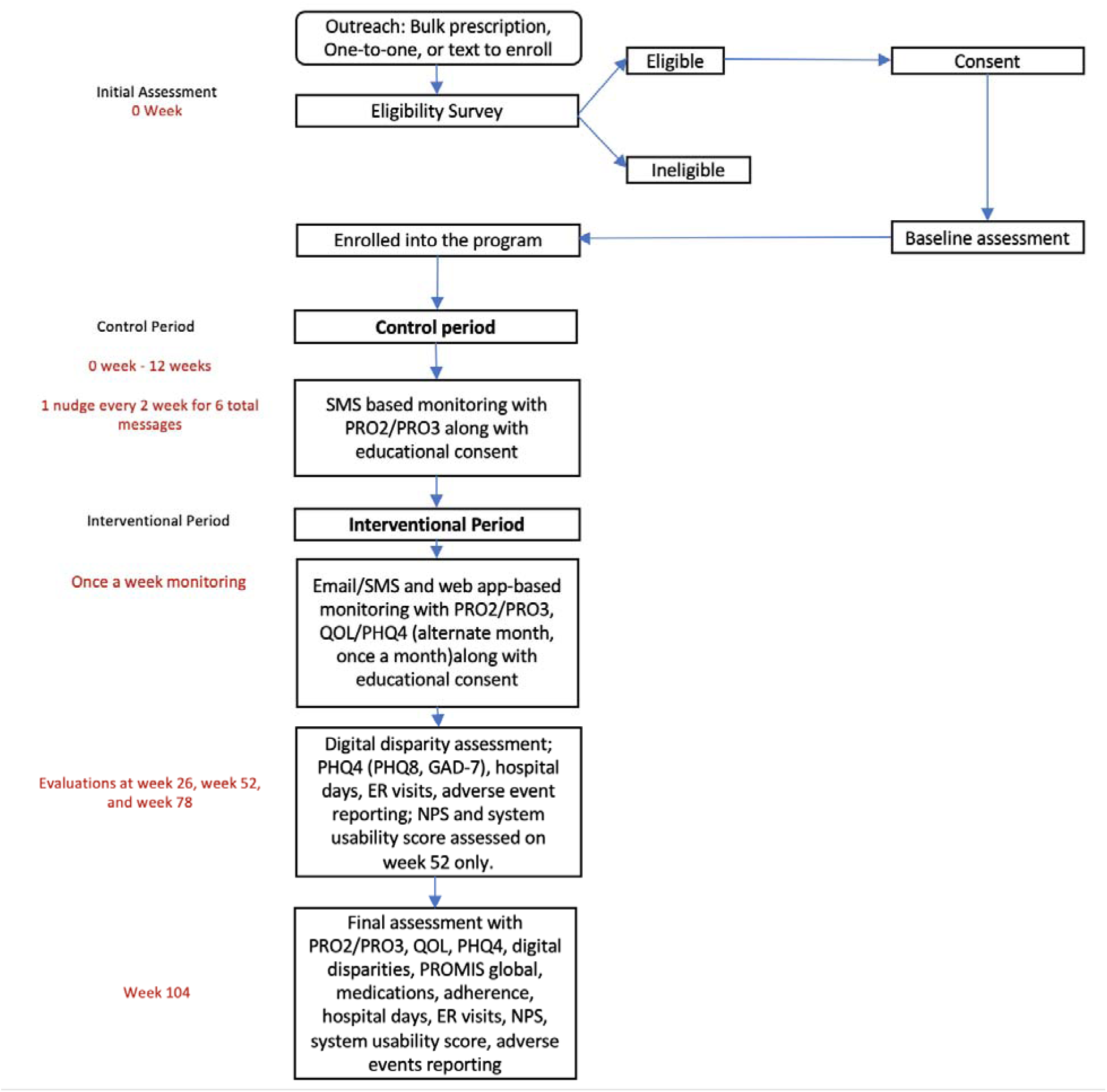
Enrollment Flow Chart. SMS – short message service, PRO2 – Patient reported outcome 2 survey, PRO3 – patient reported outcome 3 survey, QOL – quality of life, PHQ4 – Patient health questionnaire 4, PHQ8 – Patient health questionnaire 8, GAD-7 – Generalized anxiety disorder 7, ER – emergency room, NPS – net promoter score, PROMIS – Patient-Reported Outcomes Measurement Information System

### Bulk Enrollment Prescriptions

Potential research participants were identified via a computable phenotype based on IBD diagnosis (Crohn’s Disease (CD) or Ulcerative Colitis (UC)) and age (18 years or older) generated from the EHR based on each site’s specific protocol. IBD phenotype was determined by the patient’s problem list in the EHR at UC Davis Health. Mount Sinai used appointment schedules for both the IBD Center and the Infusion Center to create their prospective participant pool, as well as ICD-10 codes within their EHR. At Cleveland Clinic, patients who had an established IBD provider were pulled from a list and charts were manually reviewed for eligibility by a coordinator. Every two weeks, a data pull was generated from the EHR based on the site-specific process outlined above. This EHR data pull file included the potentially eligible patient’s name, phone number, email address, IBD diagnosis type, IBD physician, Medical Record Number, Zip code, Department code, time and date of patient’s next appointment, and myChart activity status. Using the EHR, research staff then reviewed each data pull file to confirm all of a patient’s diagnoses and screened for exclusion criteria based on study protocol. After it was reviewed, this EHR data pull file was uploaded to Prescription Universe. The Prescription Universe then served as a functional platform for enrollment, monitoring, and later intervention. The data could then be utilized for systemic outreach to patients for bulk enrollment. Once the Bulk Prescription files were uploaded to that platform, a profile for each patient listed in the file was created. Similarly, a patient profile could be created on the platform manually by the coordinator via the other two recruitment methods (i.e. if engaged by One-to-one prescription) or by a patient who scanned the QR code or texted the number on the flyers (i.e. Text to enroll prescription). Once these profiles were created within the platform, automated messages were sent to the patient either inviting them to enroll, presenting them with a link to complete the baseline assessment, or sending them messages during both the control and interventional periods. Bulk prescriptions refer to individual optional invitations to participate in research. As mentioned above, UC Davis Health and Mount Sinai utilized Prescription Universe for the initial outreach where Cleveland Clinic utilized outreach via MyChart with a link for subsequent patient enrollment via the Prescription Universe.

Once the data pull was uploaded, all potentially eligible patients listed were sent a text message (Figure 2), email message, or a MyChart message containing a link to an informational video about the study and an eligibility survey. During this process a patient could click to speak to a coordinator if further clarification was needed.

**Figure 2.**
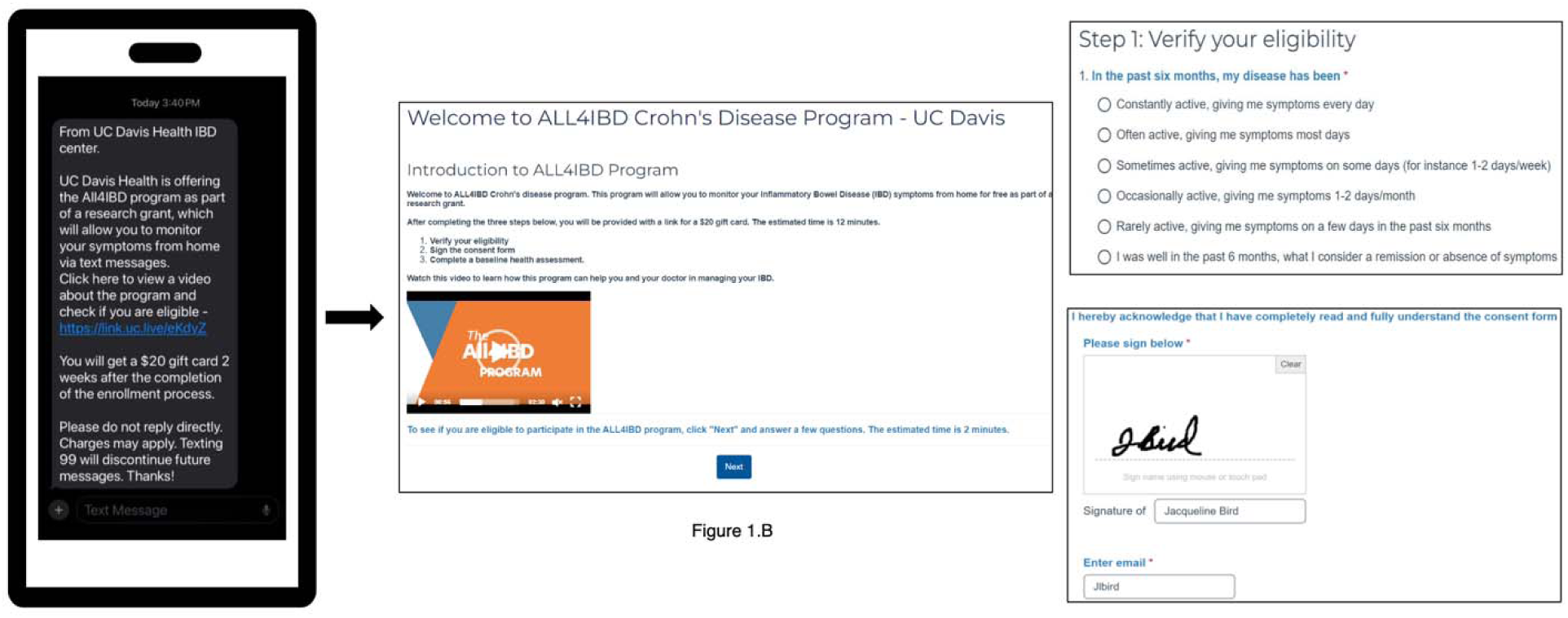
Overview of Enrollment and Outreach Process. A - SMS enrollment outreach sent to eligible patients. B - Introductory material patients see prior to enrollment. C - Eligibility survey and consent example.

Once a potential participant clicked on the link, the patient was prompted with details about the purpose of the study, study contact information, and eligibility criteria, which was determined after the patient completed a 6-question eligibility survey assessing disease activity (Figure 3).

**Figure 3.**
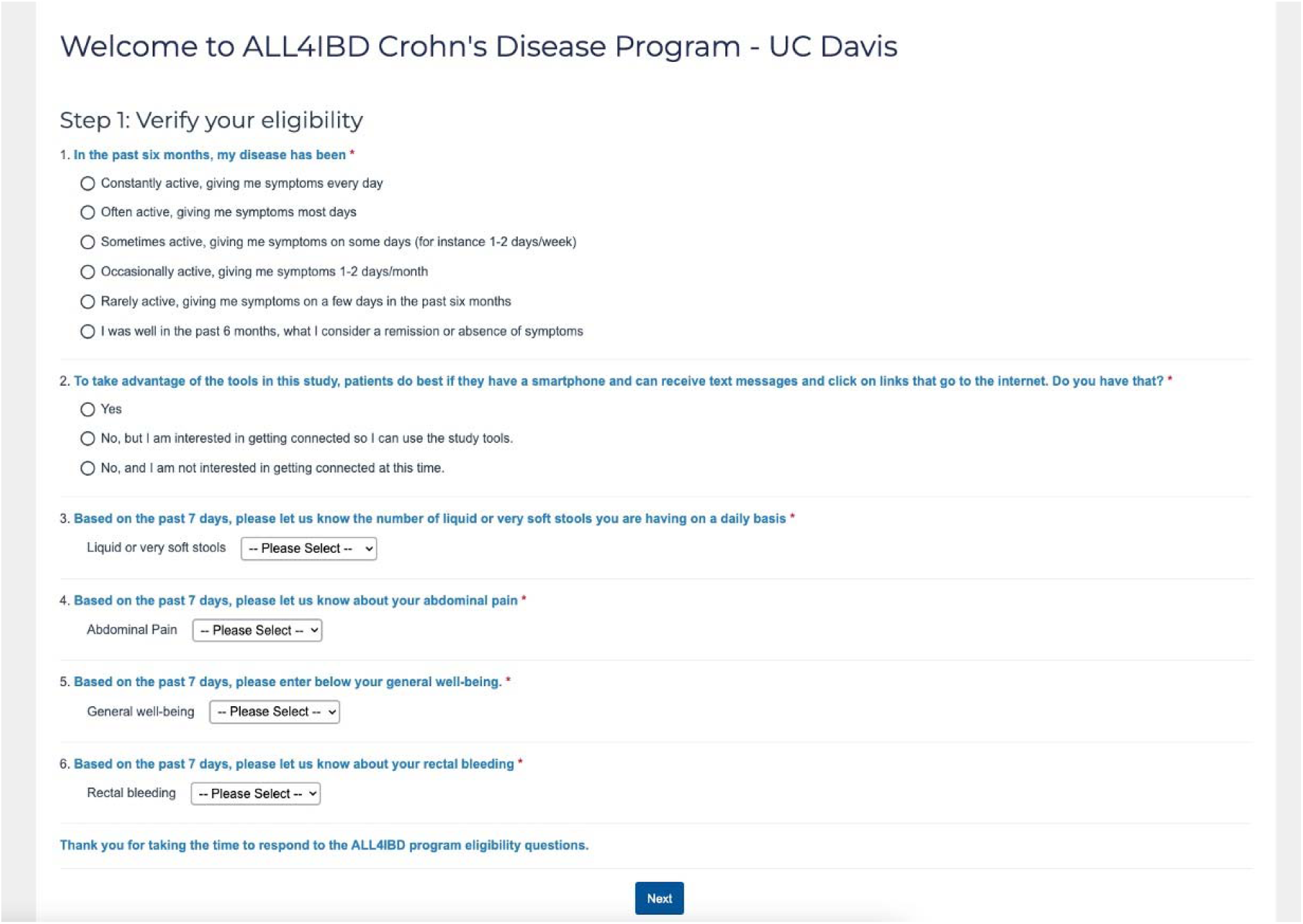
Online 6-Question eligibility questionnaire.

Subsequently, those patients interested in participating in the study were prompted to complete an electronic consent form (Figure 4).

**Figure 4.**
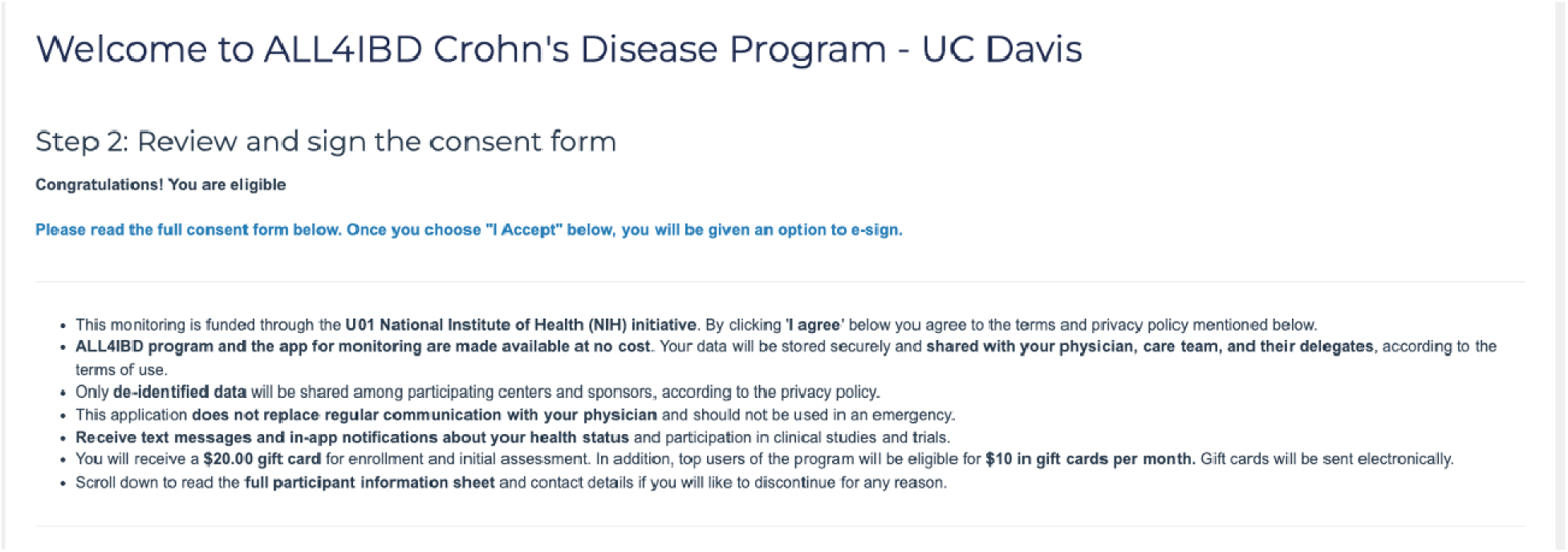
Electronic consent.

After enrollment, the patient’s baseline symptoms were assessed via virtual ePRO surveys. After the initial Bulk Prescription had occurred, the site coordinator followed-up with patients while they were on site for their scheduled clinic or treatment visit to further discuss the study and facilitate e-consenting if not already completed. Otherwise, the coordinator could follow-up via phone call and/or automated digital nudges via the Prescription Universe platform. Pre-written, automated digital “nudges” were utilized for the purpose of this study as well. A second message was queued to be sent the morning of the patient’s scheduled visit if the patient had not previously enrolled or responded to the initial bulk prescription message sent out. If there was no response after a total of three phone calls, one prescription, and one nudge (for a total of 5 contacts) the patient was deemed “lost to follow-up”. Coordinators could also manually trigger pre-written messages to send to patients that had expressed interest but requested information about the study to be re-sent digitally.

Every two weeks, a new Bulk Prescription file was generated from each site’s EHR, and the recruitment cycle was repeated. As patients continued through the trial, they were sent an educational video, and a 2-3 question survey every 2 weeks in the form of an ePRO. Patients were sent specific ePROs and educational material based on IBD phenotype.

### One-to-one Prescribing

Potential research participants who were not captured in the bulk prescription method were recruited individually using One-to-One prescriptions and text to enroll. An example of how a subject could have been missed by Bulk Prescription is that some patients may not have an IBD diagnosis listed within their problem list on the EHR but did have a diagnosis written in their outpatient clinic notes. Such patients could then be captured by one-to-one prescribing. Another example in that the one-to-one process, occurred when the coordinator retrospectively screened the previous two weeks of IBD clinic scheduled appointments. For a patient who was not already prescribed via bulk prescription and met all inclusion criteria, the coordinator had the option to manually trigger enrollment invitation through the Prescription Universe platform for that patient only. The coordinator could generate a new enrollment prescription for the patient in the form of text and email messages containing a link to the informational video and an eligibility survey. Patients interested in taking part in the study were then asked to fill out an electronic consent form and a baseline survey after providing their signature of consent and proceeded in the same path as outlined in Figure 1. Alternatively, if a coordinator was approached by a patient during their clinic visit that had not already been prescribed but expressed interest in participating and met all inclusion criteria, the same one-to-one prescription procedure described above was repeated.

### Text to enroll

Eligible patients not captured by either bulk prescription or one-to-one prescribing were also able to enroll with patient-initiated text message based activation utilizing a flyer with a QR code posted in the office at appointments. Interested patients were able to proactively text a short code that they initiated after seeing a flyer to receive further information and initiate the enrollment process. The code was posted on flyers within the IBD clinics for patients to access. Through this link, they were able to complete the onboarding module and e-consent as above and proceed through the same steps as outlined above if they met eligibility criteria on the self-reported survey.

### Intervention Phase

Providers who were taking care of enrolled patients with documented changes on ePRO assessment that fulfilled intervention criteria were notified and had the option to intervene clinically if indicated. Patients who reported worsening ePRO scores could be identified through the Prescription Universe platform. A coordinator would then message the patient’s provider via a secure EHR staff message. The results for this study do not present the completion of the intervention arm as this part of the trial is currently ongoing. The goal at this stage was to show feasibility of decentralized recruitment for a clinical trial and the outline of the ongoing ePRO process.

### Data analysis and Statistics

Data was collected then processed in Python using supporting libraries including Pandas, Numpy, and Scipy. Demographic variables including age, gender, race, smoking status, marital status, and outreach method were compared between the patients who enrolled vs did not enroll in the study assessing for statistical significance. T-test were performed for continuous variables and chi-squared test were performed for all other categorical variables. A p-value of < 0.05 was used to reflect statistical significance.

## RESULTS

### Baseline characteristics

Using three modalities including bulk outreach, one-to-one prescription, and text to enroll, we were able to approach 6,687 patients over 12 months and successfully enroll 543 patients across the three IBD referral centers (Figure 5).

**Figure 5.**
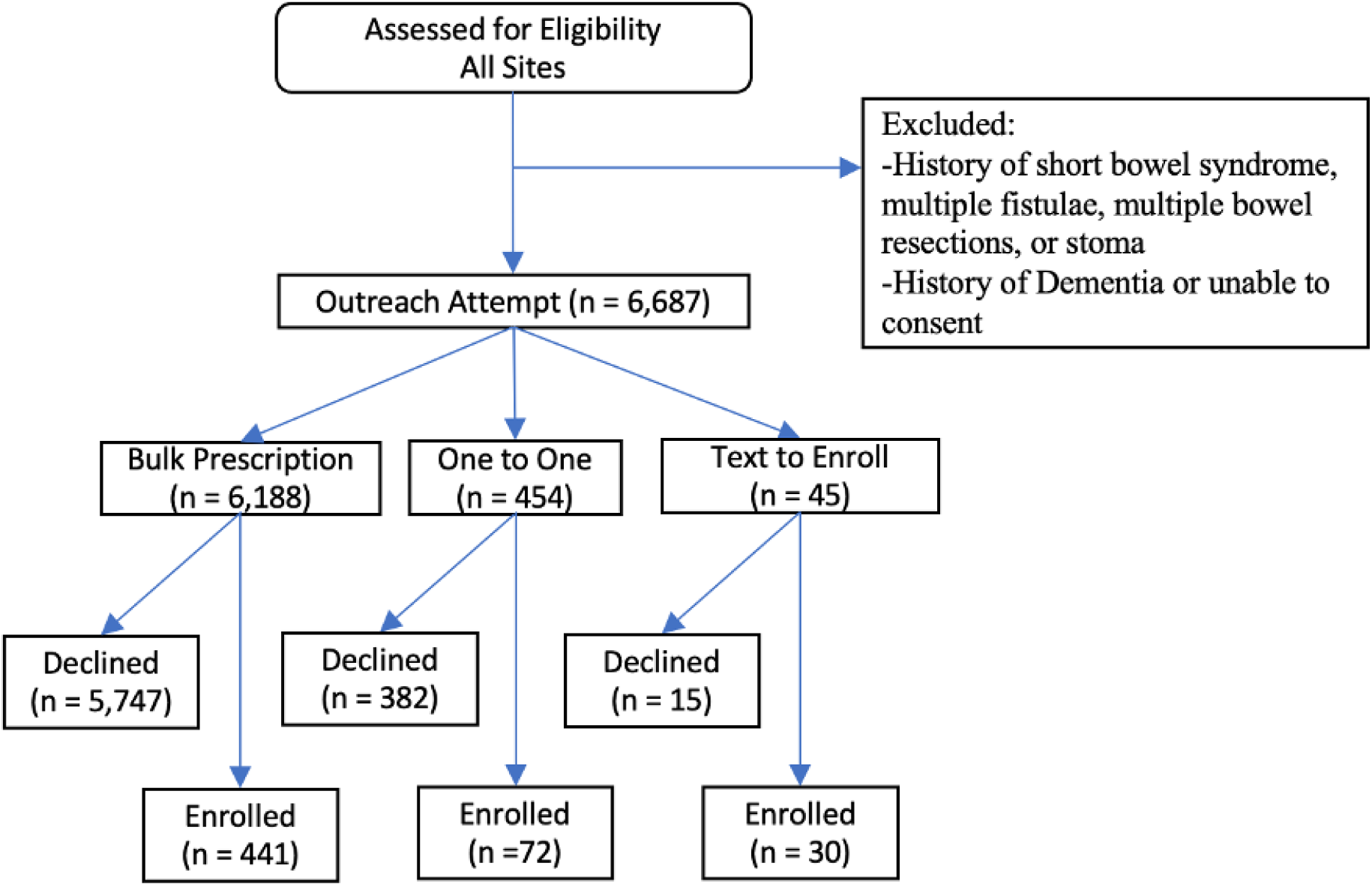
Enrollment flow chart by outreach modality.

In the combined analysis, enrolled patients tended to be middle aged (mean age 40.7 ± 15.7 years), female (63.3% vs 36.7%, p = <0.001) and White (75.3%, p = <0.001). Smoking status was not a significant variable between enrolled vs not enrolled patients. When assessing IBD phenotype, there was also no significant difference between enrolled vs not enrolled patients. Enrolled patients had similar rates of both UC compared to not enrolled (39.0% v 39.0%) and CD (60.6% vs 61.0%). Both patient groups also demonstrated similar rates of marital status 46.4% vs 46.5% in enrolled vs unenrolled. In the combined cohort, 81.2% of enrolled patients were recruited from the bulk prescription method. Gender (P<0.01), and race (P<0.01) were statistically significant in determining digital clinical trial enrollment and engagement (Table 1).

**Table 1.** All Clinical Sites Combined Data.

| Variable |  | Not Enrolled |  | Enrolled |  | P – Value |
| --- | --- | --- | --- | --- | --- | --- |
|  |  | N=6144 | % | N=543 | % |  |
| Age |  | 41.3 +- 16.92 |  | 40.7 +- 15.65 |  | 0.532 |
| Gender | Female | 3021 | 49.17 | 344 | 63.35 | <0.001 |
|  | Male | 3119 | 50.76 | 199 | 36.65 |  |
|  | Unknown | 4 | 0.07 | 0 | 0.00 |  |
| Race | White | 4532 | 73.76 | 409 | 75.32 | <0.001 |
|  | Asian | 247 | 4.02 | 20 | 3.68 |  |
|  | Black or | 429 | 6.98 | 34 | 6.26 |  |
|  | African |  |  |  |  |  |
|  | American |  |  |  |  |  |
|  | Hispanic | 92 | 1.50 | 7 | 1.29 |  |
|  | Other | 558 | 9.08 | 45 | 8.29 |  |
|  | Unknown | 286 | 4.65 | 28 | 5.16 |  |
| IBD | Crohn's disease | 3749 | 61.02 | 329 | 60.59 | 0.935 |
| Phenotype | Ulcerative colitis | 2395 | 38.98 | 214 | 39.41 |  |
| Smoking | Former Smoker | 995 | 16.19 | 79 | 14.55 | 0.421 |
| Status | Heavy or | 175 | 2.85 | 19 | 3.50 |  |
|  | Everyday |  |  |  |  |  |

| **Smoker** |  |  |  |  |  |  |
| --- | --- | --- | --- | --- | --- | --- |
|  | **Light Smoker** | **112** | ***1.82*** | **15** | ***2.76*** |  |
|  | **Never** | **3782** | ***61.56*** | **335** | ***61.69*** |  |
|  | **Unknown** | **1080** | ***17.58*** | **95** | ***17.50*** |  |
| **Marital** | **Married/ Life** | **2860** | ***46.55*** | **252** | ***46.41*** | **0.453** |
| **Status** | **Partner** |  |  |  |  |  |
|  | **Separated or** | **262** | ***4.26*** | **31** | ***5.71*** |  |
|  | **divorced** |  |  |  |  |  |
|  | **Single** | **2632** | ***42.84*** | **232** | ***42.73*** |  |
|  | **Unknown/Other** | **306** | ***4.98*** | **21** | ***3.87*** |  |
|  | **Widowed** | **84** | ***1.37*** | **7** | ***1.29*** |  |
| **Outreach** | **Bulk** | **5747** | ***93.54*** | **441** | ***81.22*** | **<0.001** |
|  | **One to One** | **382** | ***6.22*** | **72** | ***13.26*** |  |
|  | **Text to Enroll** | **15** | ***0.24*** | **30** | ***5.52*** |  |

Individual site data is provided in Supplementary Tables 1-3. At UC Davis Health (Supplementary Table 1), 549 patients were approached over 2 months and 70 patients were successfully enrolled. Enrolled patients were also middle aged (mean age 45.2 ± 17.2), female (61.4%) and White (74.2% vs 58.6%, p=0.035). At Cleveland Clinic (Supplementary Table 2), 1,746 patients were approached, and 133 patients were enrolled. Enrolled patients were younger (mean age 44.0 ± 14.2 vs 49.1 ± 17.0, p= <0.001), and predominantly female (74.4% vs 50.8%, p=<0.001). Both enrolled and not enrolled patients were predominately White (85.0% vs 84.5%). At the lead site, Mount Sinai (Supplementary Table 3), 4,392 patients were approached, and 340 patients were enrolled. Enrolled patients were middle aged (mean age 41.3 ± 16.9) and White (71.3% vs 71.8%).

## DISCUSSION

Technological advancements now offer an exciting opportunity for improved clinical recruitment and patient assessment. Our study is among the first to employ an automated digital outreach program for recruitment and monitoring in a multicenter pragmatic clinical trial in IBD. In this first stage of the trial, we sought to demonstrate feasibility of multicenter decentralized recruitment with ongoing evaluation of ePROs in patients with IBD. We found that by utilizing a combination of various engagement methods with human in the loop strategy we could create a feasible decentralized strategy for clinical trial recruitment. Bulk prescription allowed for the greatest outreach to patients while maximizing resource efficiency. Ongoing assessments via ePROs could then continue in a decentralized manner for the remainder of the study.

In 2013, the STRIDE program was created by the International Organization for the Study of Inflammatory Bowel Disease (IOIBD) and put forth expert consensus guidelines for selecting goals in treat-to-target strategies.^5^ In both UC and CD, PROs of remission serve as an essential goal in the treat-to-target strategy. These guidelines were further updated with the STRIDE-II initiative in 2021.^6^ STRIDE-II reaffirmed the importance of PROs as a standard clinical target that should be assessed early and often throughout the disease course. With these ideas in mind, decentralized monitoring of ePROs offer an exciting opportunity to more accurately assess a patient’s clinical course. Patients monitored remotely can be assessed more frequently than in person visits would allow through an online platform accessible via their electronic device. This data can then be used in a modern treat-to-target approach to identify patients who are either at risk or in the early stages of clinical deterioration. The end goal of the decentralized ePRO monitoring is then to be able to identify those patients who are at the earliest possible stage of clinical deterioration and intervene before they decompensate. This process begins though with the decentralized electronic recruitment.

Using the Prescription Universe platform, we were able to automate much of the enrollment process to include not only the physical outreach through text and email but also the education and consent. This is a great improvement from the historic clinical trial recruitment procedure. Traditionally, patients are both recruited to participate in a study and evaluated for progress during their appointment with a physician. This places significant time constraints on the number of patients able to be recruited as well as how frequently they can be assessed during a trial with standard in person PRO. Decentralization helps to maximize the team’s efficiency to recruit patients as well as monitor them more frequently with ePRO. Monitoring with ePROs can then identify patients in need earlier and more frequently than in person visits. Decentralization thus optimizes both the recruitment and intervention efforts of a clinical trial allowing for a greater chance of completion.

We also demonstrated the feasibility of applying a decentralized strategy across multiple institutions. Our study took place at three different academic centers across the United States. At each center, we were able to apply a decentralized strategy and have adequate accrual for our clinical trial while maintaining compliance with a centralized IRB. Coordinating a multicenter study also poses its own set of intrinsic limitations. With the addition of each center to a clinical trial, the resources required, and administrative oversight increase dramatically. Adhering with multiple IRBs can delay study implementation, rightfully so for patient safety. Additionally, once a study has begun, combining patient data that has been extracted by various teams also presents another potential source of error at multiple steps. We addressed these issues by establishing a close working relationship between the administration and investigators at each site. We coordinated multi-institutional biweekly meetings with each center’s principal investigators as well as research coordinators. A common protocol was created which was submitted to Advarra IRB. This allowed us to have standardized protocols taking place at each site. Additionally, the data was reviewed by each site prior to submission to a central data analyst who handled the combining and analysis.

Further limitations included outreach to patients in very remote areas without internet access or older patients who lacked technical computer skills. For example, patients who live in areas without internet access would have too much difficulty continuing to participate. We also found other groups for religious reasons did not always have access to internet which may bias their responses. Instances such as these could explain the significant study results regarding gender and race. The initial eligibility survey asked patients whether they had digital access/literacy, and we did not capture many patients stating that they did not. This could be because the initial means of communication was digital, so those who were not connected were mostly excluded to begin with. Additionally, we utilized only Epic for the prescreening process given this was the EHR already in place at each site however this process could be implemented universally for prescreening with other EHRs then uploading directly to the Commure Database. Lastly, the only language used throughout the study materials was English and this could further contribute to our results.

Our findings align with previous studies that have utilized decentralized recruitment methods and shown feasibility to clinical trial accrual. Though we did not compare our findings to traditional recruitment strategies specifically within this study, others have not only confirmed feasibility of decentralize recruitment but also superiority to traditional methods. Sommer et al. evaluated the efficacy of decentralized recruitment through virtual visits compared to traditional methods and found a higher recruitment rate in the decentralized approach compared to the conventional method.^7^ Similar results indicating the superiority over traditional approaches at clinical trial accrual were also observed in studies assessing decentralized recruitment through virtual methods in patients with irritable bowel syndrome and COVID-19.^8–10^ Furthermore, decentralized recruitment and monitoring has also demonstrated feasibility when attempting to reach patients residing in rural areas.^11–13^ This not only addresses a major obstacle contributing to clinical trial failures but also offers cost-effectiveness and improved outreach particularly towards at risk patient populations who may not be able to attend in person frequent visits.

The need for improving clinical trial performance is great. Each clinical trial failure represents poor stewardship of public funds and a waste of scarce resources. Clinical trials then must adapt to account for this need in both the recruitment phase as well as the interventional phase. Innovation can provide greater access to clinical trials to both ensure their completion and maximize their performance. This is one of the first studies to show feasibility and successful recruitment of patients with IBD using automated digital outreach. We found a combination of different outreach and engagement methods (bulk recruitment, one-to-on, and text to enroll) with human in the loop as the most effective strategy for clinical trial accrual. These strategies highlight the potential to implement decentralized digital recruitment and monitoring with ePROs in clinical trials for other chronic diseases going forward. Future studies evaluating the efficacy of decentralized recruitment and the optimization of existing protocols will help to solidify this method as a standard option in clinical trial planning.

## Data Availability

All data produced in the present study are available upon reasonable request to the authors

---

**Supplementary Table 1.** UC Davis Site Data. Comparative Analysis between Enrolled vs Not Enrolled Patients

Comparative Analysis between Enrolled vs Not Enrolled Patients
| Variable |  | Not Enrolled |  | Enrolled |  | P – Value |
| --- | --- | --- | --- | --- | --- | --- |
|  |  | N = 479 | % | N = 70 | % |  |
| Age |  | 48.95 +- 18.11 |  | 45.2 +- 17.15 |  | 0.111 |
| Gender | Female | 251 | 52.40 | 43 | 61.43 | 0.341 |
|  | Male | 228 | 47.60 | 27 | 38.57 |  |
| Race | White | 281 | 58.66 | 52 | 74.29 | 0.035 |
|  | Asian | 35 | 7.31 | 2 | 2.86 |  |
|  | Black or African American | 39 | 8.14 | 1 | 1.43 |  |
|  | Hispanic | 34 | 7.10 | 3 | 4.29 |  |
|  | Other | 36 | 7.52 | 4 | 5.71 |  |
|  | Unknown | 54 | 11.27 | 8 | 11.43 |  |
| IBD Phenotype | Crohn's disease | 255 | 53.24 | 45 | 64.29 | 0.206 |
|  | Ulcerative colitis | 224 | 46.76 | 25 | 35.71 |  |
| Smoking Status | Former Smoker | 134 | 27.97 | 14 | 20.00 | 0.192 |
|  | Heavy or Everyday Smoker | 21 | 4.38 | 3 | 4.29 |  |
|  | Light Smoker | 5 | 1.04 | 3 | 4.29 |  |
|  | Never | 311 | 64.93 | 49 | 70.00 |  |
|  | Unknown | 8 | 1.67 | 1 | 1.43 |  |
| Marital Status | Married/ Life Partner | 230 | 48.02 | 36 | 51.43 | 0.343 |
|  | **Separated or Divorced** | **28** | **5.85** | **6** | **8.57** |  |
|  | **Single** | **174** | **36.53** | **26** | **37.14** |  |
|  | **Unknown/Other** | **33** | **6.89** | **2** | **2.86** |  |
|  | **Widowed** | **14** | **2.92** |  | **0.00** |  |
| **Outreach** | **Bulk** | **282** | **58.87** | **42** | **60.00** | **0.858** |
|  | **One to One** | **197** | **41.13** | **28** | **40.00** |  |
|  | **Text to Enroll** | **0** | **0.00** | **0** | **0.00** |  |

**Supplementary Table 2.** Cleveland Clinic Site Data. Comparative Analysis between Enrolled vs Not Enrolled Patients

Comparative Analysis between Enrolled vs Not Enrolled Patients
| Variable |  | Not Enrolled |  | Enrolled |  | P –<br>Value |
| --- | --- | --- | --- | --- | --- | --- |
|  |  | N = | % | N = | % |  |
|  |  | 1613 |  | 133 |  |  |
| Age |  | 49.09 +- 16.99 |  | 43.99 +- 14.20 |  | <0.001 |
| Gender | Female | 820 | 50.84 | 99 | 74.44 | <0.001 |
|  | Male | 793 | 49.16 | 34 | 25.56 |  |
| Race | White | 1363 | 84.50 | 113 | 84.96 | 0.545 |
|  | Asian | 21 | 1.30 | 2 | 1.50 |  |
|  | Black or African American | 143 | 8.87 | 9 | 6.77 |  |
|  | Hispanic | 20 | 1.24 | 2 | 1.50 |  |
|  | Other | 17 | 1.05 |  | 0.00 |  |
|  | Unknown | 49 | 3.04 | 7 | 5.26 |  |
| IBD Phenotype | Crohn's disease | 978 | 60.52 | 83 | 62.41 | 0.66 |
|  | Ulcerative colitis | 635 | 39.48 | 50 | 37.59 |  |
| Smoking | Former Smoker | 432 | 26.78 | 34 | 25.56 | 0.066 |
| Status | Heavy or Everyday Smoker | 76 | 4.71 | 10 | 7.52 |  |
|  | Light Smoker | 30 | 1.86 | 4 | 3.01 |  |
|  | Never | 1071 | 66.40 | 83 | 62.41 |  |
|  | **Unknown** | **4** | ***0.25*** | **2** | ***1.50*** |  |
| **Marital Status** | **Married/ Life Partner** | **909** | ***56.35*** | **70** | ***52.63*** | **0.864** |
|  | **Separated or Divorced** | **114** | ***7.07*** | **11** | ***8.27*** |  |
|  | **Single** | **544** | ***33.73*** | **48** | ***36.09*** |  |
|  | **Unknown/Other** | **15** | ***0.93*** | **2** | ***1.50*** |  |
|  | **Widowed** | **31** | ***1.92*** | **2** | ***1.50*** |  |
| **Outreach** | **Bulk** | **1605** | ***99.50*** | **109** | ***81.95*** | **<0.001** |
|  | **One to One** | **0** | ***0.00*** | **0** | ***0.00*** |  |
|  | **Text to Enroll** | **8** | ***0.50*** | **24** | ***18.05*** |  |

**Supplementary Table 3.** Mount Sinai Site Data. Comparative Analysis between Enrolled vs Not Enrolled Patients

Comparative Analysis between Enrolled vs Not Enrolled Patients
| Variable |  | Not Enrolled |  | Enrolled |  | P – Value |
| --- | --- | --- | --- | --- | --- | --- |
|  |  | N= | % | N = | % |  |
|  |  | 4052 |  | 340 |  |  |
| Age |  | 40.7 +- 15.65 |  | 41.3 +- 16.92 |  | 0.532 |
| Gender | Female | 1950 | 48.12 | 202 | 59.41 | 0.001 |
|  | Male | 2098 | 51.78 | 138 | 40.59 |  |
|  | Unknown | 4 | 0.10 | 0 | 0.00 |  |
| Race | White | 2888 | 71.27 | 244 | 71.76 | 0.937 |
|  | Asian | 191 | 4.71 | 16 | 4.71 |  |
|  | Black or African American | 247 | 6.10 | 24 | 7.06 |  |
|  | Hispanic | 38 | 0.94 | 2 | 0.59 |  |
|  | Other | 505 | 12.46 | 41 | 12.06 |  |
|  | Unknown | 183 | 4.52 | 13 | 3.82 |  |
| IBD Phenotype | Crohn's disease | 2516 | 62.10 | 201 | 59.12 | 0.006 |
|  | Ulcerative colitis | 1536 | 37.91 | 139 | 40.88 |  |
| Smoking Status | Former Smoker | 429 | 10.59 | 31 | 9.12 | 0.896 |
|  | Heavy or Everyday Smoker | 78 | 1.92 | 6 | 1.76 |  |
|  | Light Smoker | 77 | 1.90 | 8 | 2.35 |  |
|  | **Never** | **2400** | **59.23** | **203** | **59.71** |  |
|  | **Unknown** | **1068** | **26.36** | **92** | **27.06** |  |
| **Marital Status** | **Married/ Life Partner** | **1721** | **42.47** | **146** | **42.94** | **0.529** |
|  | **Separated or divorced** | **120** | **2.96** | **14** | **4.12** |  |
|  | **Single** | **1914** | **47.24** | **158** | **46.47** |  |
|  | **Unknown/Other** | **258** | **6.37** | **17** | **5.00** |  |
|  | **Widowed** | **39** | **0.96** | **5** | **1.47** |  |
| **Outreach** | **Bulk** | **3860** | **95.26** | **290** | **85.30** | **<0.001** |
|  | **One to One** | **185** | **4.57** | **44** | **13.00** |  |
|  | **Text to Enroll** | **7** | **0.17** | **6** | **1.70** |  |

